# *In silico* perturbations provide multivariate interpretability in predicting post-lung transplant outcomes

**DOI:** 10.1101/2024.10.19.24315817

**Authors:** Lucy Luo, Marcin Możejko, Nikolay S. Markov, Alec Peltekian, Suror Mohsin, Mary Carns, Phillip Cooper, Jeffrey Lysne, Anthony Joudi, Alan Betensley, Bradford C. Bemiss, Catherine Myers, Ankit Bharat, Rade Tomic, Ambalavanan Arunachalam, Ewa Szczurek, GR Scott Budinger, Alexander V. Misharin, Mrinalini Venkata Subramani

**Affiliations:** Northwestern University Feinberg School of Medicine, Division of Pulmonary and Critical Care Medicine, Department of Medicine, Simpson Querrey Lung Institute for Translational Sciences. Chicago, IL, USA; University of Warsaw, Faculty of Mathematics, Informatics and Mechanics. Warsaw, Poland; Northwestern University McCormick School of Engineering and Applied Science, Department of Computer Science. Chicago, IL, USA; Northwestern Memorial Hospital. Chicago, IL, USA; Northwestern University Feinberg School of Medicine, Department of Surgery (Thoracic Surgery). Chicago, IL, USA; Helmholtz Zentrum Munich, Institute for AI for Health. Munich, Germany

## Abstract

Lung transplantation is a life-saving therapy for end-stage lung disease but has the poorest survival among solid organ transplants. We analyzed standardized electronic health record (EHR) data from the United Network for Organ Sharing (UNOS) to predict one-, three-, and five-year survival and favorable long-term outcomes post-lung transplant. We applied two multivariate machine learning approaches, XGBoost or a tabular BERT model called EHRFormer, to data from 43,869 first-time lung transplant recipients (1987–2022). XGBoost and EHRFormer identified features that align closely with established risk factors for worse outcomes such as length of index stay, recipient age, and creatinine at the time of transplant. We developed a simple perturbation method with EHRFormer to probe *in silico* multivariate interactions between features that influence model prediction. Despite their attention to known risk factors, machine learning applied to EHR data collected by UNOS poorly predict one-, three-, and five-year mortality after lung transplant.

## Introduction

Lung transplantation is the only viable treatment option that improves survival and quality of life for patients with advanced lung disease and respiratory failure. Despite improvements in surgical techniques, immunosuppressive strategies, perioperative management, supportive strategies, and approaches for donor lung allocation over the years, lung transplantation is persistently associated with poor survival relative to other solid organ transplants, with a median survival of 5.8 years (1990-2014) ^2^ and a mean survival of 9.28 years. ^1^ Approximately 10-15% of all deaths after lung transplant occur in the first year. ^3^ In patients who survived one year after transplant, median survival is 10.2 years. ^4^ As a result, one-year mortality is a trajectory-defining event and is the focus of public reporting of transplant outcomes. Recently, investigators have suggested that three- or five-year outcomes provide additional data with respect to center-specific transplant outcomes, leading some to suggest these outcomes be publicly reported ^5^ . We reasoned that prediction of one-, three- and five-year survival based on factors available early in the transplant course could identify patients for targeted interventions. Further, we reasoned that waitlist and peri-transplant factors in the donor and recipient that predict outcomes might include modifiable factors to improve outcomes. Conventional analyses of lung transplant outcomes use univariate or multivariate statistical methods on pre-selected variables. As such, available predictors of lung transplant outcomes vary by center and perform poorly. ^4,6–15^ Machine learning is a powerful approach to identify predictors of outcomes from clinical data, leading to its growing use in clinical research and care. However, its application to EHR data collected by the United Network for Organ Sharing has yielded disappointing results.^16,17^ As machine learning approaches have dramatically improved since the publication of those studies, we aimed to test whether modern machine learning methods could predict one-, three-, and five-year lung transplant outcomes using data from the UNOS database. We additionally sought to leverage model interpretability, which a significant number of sophisticated machine learning and deep learning approaches lack, to validate our findings. By applying XGBoost^18^ and a Bidirectional Encoder Representations from Transformers (BERT)^19^ -based model, EHRformer, we validated many established risk factors for one year mortality. Although BERT-based models like EHRFormer do not explicitly provide information about the factors that drive their predictions, we developed a perturbation method in EHRFormer that investigates the hypothetical effect of changing multiple variables simultaneously. In terms of performance, both weakly predict one-, three-, and five-year mortality and only modestly better when predicting lung function at one year. Performance worsened when index stay data were excluded. Our findings suggest that UNOS data, even with state-of-the-art machine learning models, poorly predict lung transplant outcomes.

## Results

### The UNOS dataset reveals changes in lung transplant practices and outcomes in the US over time

The UNOS dataset is a national, standardized database of clinical and demographic information for lung transplant candidates. ^20^ For this study, we focused on first-time lung transplant recipients aged >18 years. Because the UNOS dataset is cumulative, it reflects overall changes in national lung transplant practice over time. Our exploratory analysis of these patients suggested these changes in practice explain a significant amount of variance in the dataset (Figure 1A, 1B). For example, the proportion of transplants performed for restrictive lung disease has increased relative to those performed for obstructive lung disease. Additionally, ischemic time, age, and FEV1% at transplant have increased over time, reflecting changes in organ storage and allocation, recipient characteristics, and indication type.

**Figure 1:**
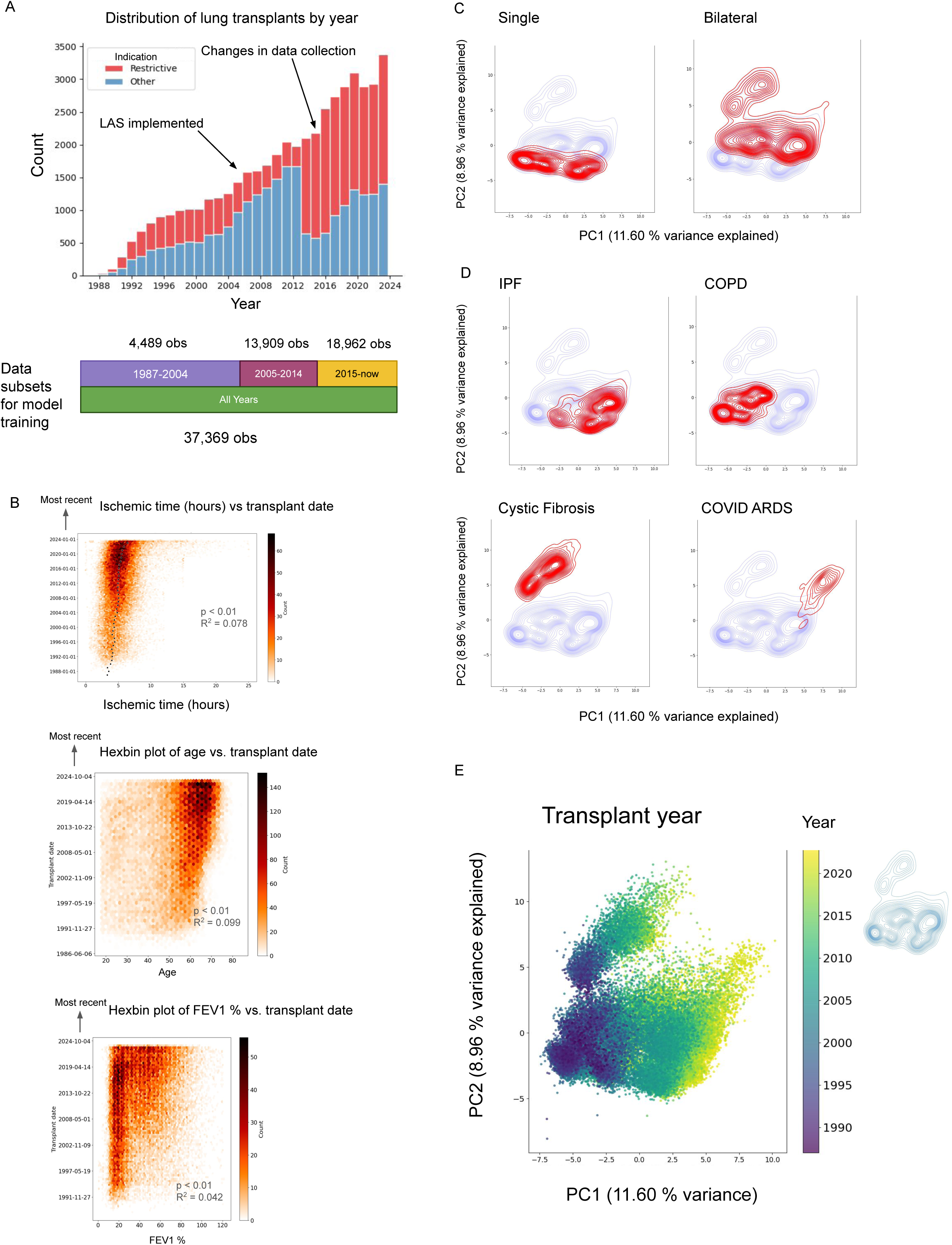
An exploratory analysis of the UNOS dataset reveals distinct indication-drive groupings and time-associated trends. A. Histogram of the number of lung transplants by year since 1987 split by restrictive indication (red). The timeline schematic below the x axis indicates how many observations were included for modeling for each model across various time frames. B. Hexbin plots showing from top to bottom the relationship of ischemic time (hours), recipient age, and FEV1% at the time of transplant with sorted transplant date. Black dots on the ischemic time plot indicate the yearly median of ischemic time. P values and R-squared values were derived from linear regression on all three variables vs transplant year. C. Principal components contour plots split by single and bilateral transplants, indicated in red. D. Principal components contour plots split by indication (red). E. Principal components scatterplot colored by transplant year with reference contour plot on the right. Early to most recent transplant years are illustrated from dark to light respectively.

An initial exploration of all relevant waitlist, peri-transplant, and follow-up features (at one, three, and five years) yielded 780 features with at least 1 observation (Table S1, Figure S1A). There were distinct patterns of feature missingness (Figure S1B), coinciding with the introduction of the Lung Allocation Score (LAS) in May 2005 ^21^ and changes in data collection introduced in 2015 (Figure 1A, S1B). Accordingly, we performed modeling separately for each of these three time periods using all features available within a given period. For the entire time period, we used only shared features. To guide feature selection for machine learning, we performed principal components analysis to identify influential features and trends. We included 47,864 first-time lung transplant recipients >18 years old and 367 features with ≤10% missing data (Figure S1C, Table S1), covering waitlist, peri-transplant, and follow-up features at one, three, and five years. Lung transplant type, indication, lung function, and transplant year explain substantial variation in the dataset (Figure 1C-E).

### XGBoost and EHRFormer fail to predict one year mortality

We ran 4 separate models based on waitlist and peri-transplant features from the time frames illustrated in Figure 1A and S1B. Features with at least 90% completeness within each time frame were included, and for the model covering all years, only features with ≥90% completeness across all years were used. The number of features in each time period is shown in Figure 2A. A data dictionary for these features is provided in Table S1. To address class imbalance, we applied downsampling to achieve a 1:1 ratio for each outcome. Additionally, individuals who died within 90 days of the index stay were excluded from mortality prediction and analyzed separately to prevent index stay length from biasing the model. A bar graph visualizing the class imbalance of outcomes is shown in Figure 2B. Final test AUCs were uniformly poor, ranging from 0.58 to 0.64 for EHRFormer (0.64 for all years) and 0.62 to 0.63 for XGBoost (0.63 for all years) (Figures 2C, S2A, Table 1). Confusion test set matrices for both models are shown in Figure S2B. Figure 2D shows test AUROC, accuracy, F1, precision, recall, and specificity for all models.

**Figure 2:**
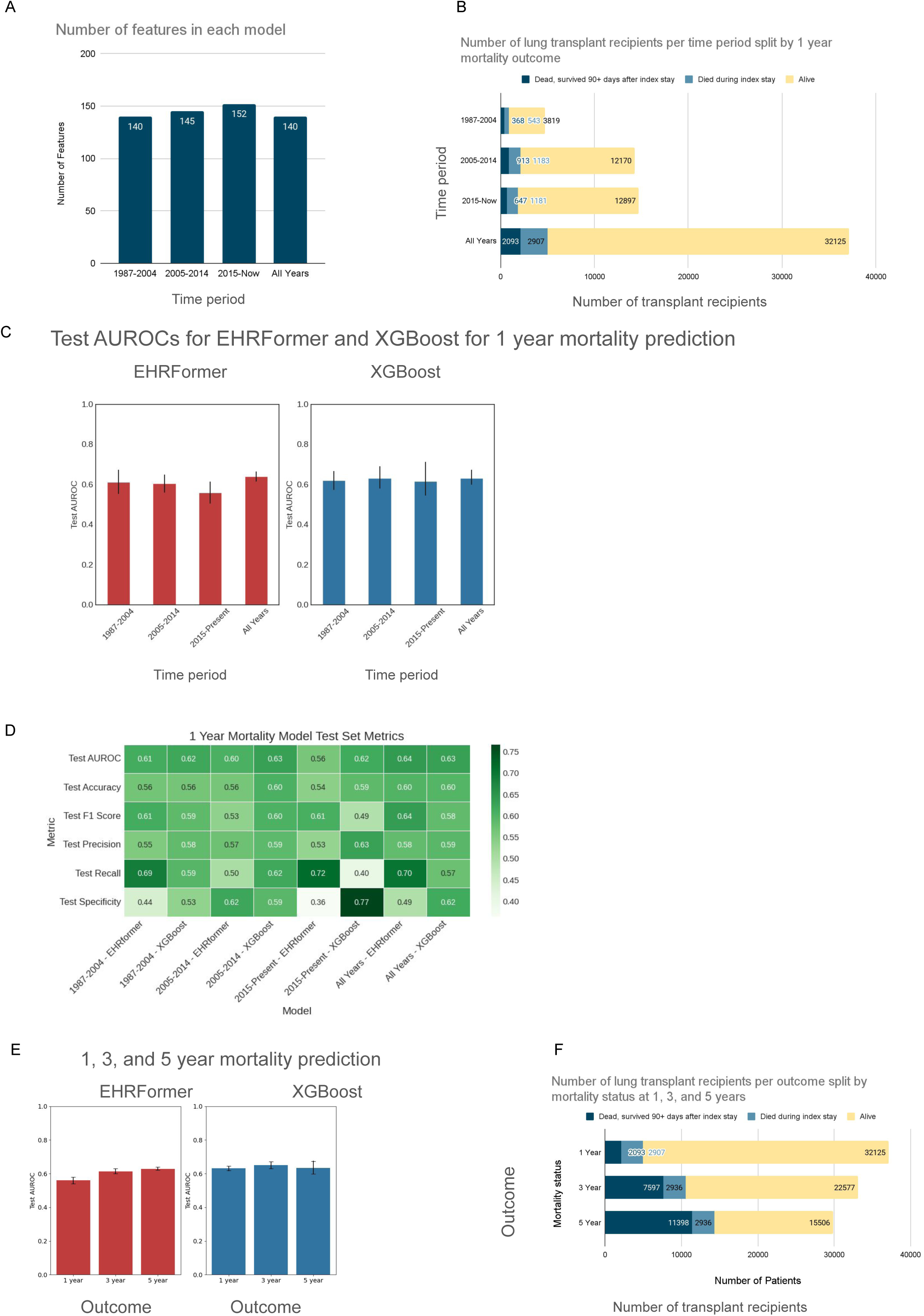
EHRFormer and XGBoost predict one-, three-, and five-year mortality with modest performance. A. The number of features used in each of the 4 different models. A data dictionary of these features is available in Table S1. B. The number of patients in each model split by class outcome of 1 year mortality. Teal indicates those who were excluded for modeling if they died within 90 days of their index stay. D. Test set AUROCs for EHRFormer (red) and XGBoost (blue) prediction of 1 year mortality across the 4 models. Error bars indicate 95% CIs based on 50 bootstraps of the test set. D. Heatmap of all test set metrics normalized within each metric (row-wise) including AUROC, accuracy, F1 score, precision, recall, and specificity for EHRFormer and XGBoost across the 4 models. E. Test set AUROCs for EHRFormer (red) and XGBoost (blue) prediction of 1-, 3-, and 5-year mortality. Error bars indicate 95% CIs based on 50 bootstraps of the test set. F. The number of patients split by class outcome of 1-, 3-, and 5-year mortality. Teal indicates those who were excluded for modeling if they died within 90 days of their index stay.

**Table 1:** Test AUROCs for EHRFormer and XGBoost predicting one year mortality across key time periods in the UNOS dataset.

| Model | 1987-2004 | 2005-2014 | 2015-now | All years |
| --- | --- | --- | --- | --- |
| EHRFormer | 0.61 [0.56,0.67]<br>95% CI | 0.60 [0.56,0.65]<br>95% CI | 0.56 [0.50,0.62]<br>95% CI | 0.64 [0.61, 0.66] 95%<br>CI |
| XGBoost | 0.62 [0.55,0.72]<br>95% CI | 0.63 [0.58,0.69]<br>95% CI | 0.62 [0.58,0.66]<br>95% CI | 0.63 [0.60, 0.67] 95%<br>CI |

**Table 2:** Test AUROCs for EHRFormer and XGBoost predicting mortality at one, three-, and five-years post-transplant.

| Model | 1-year test AUROC | 3-year test AUROC | 5-year test AUROC |
| --- | --- | --- | --- |
| EHRFormer | 0.64 [0.61, 0.66] 95% CI | 0.61 [0.60, 0.63] 95% CI | 0.63 [0.62, 0.64] 95% CI |
| XGBoost | 0.63 [0.60, 0.67] 95% CI | 0.65 [0.63, 0.67] 95% CI | 0.63 [0.62, 0.64] 95% CI |

### XGBoost and EHRFormer fail to predict one-, three-, and five-year mortality

XGBoost and EHRFormer performed poorly in predicting one-, three-, and five-year mortality, with test AUROCs ranging from 0.61 to 0.65 across all tasks and models (Figure 2E). Class imbalance in the positive and negative groups for these tasks was addressed through downsampling (Figure 2F). Model performance remained consistent across the one-, three-, and five-year mortality predictions (Figure S3).

### XGBoost and EHRFormer modestly predict patients with poor lung function at one year

Using the same waitlist and peri-transplant features (≤10% missing) from the mortality models, we predicted 1-year lung function after downsampling for class imbalance, resulting in four models (Figure 2A, Table S1, and Figure 3A). EHRFormer and XGBoost modestly predicted 1-year lung function with final test AUCs ranging from 0.72 to 0.74 for EHRFormer (0.74 for all years) and 0.74 to 0.79 for XGBoost (0.76 for all years) (Figure 3B, 3C, S3A, S3B, and Table 3).

**Figure 3:**
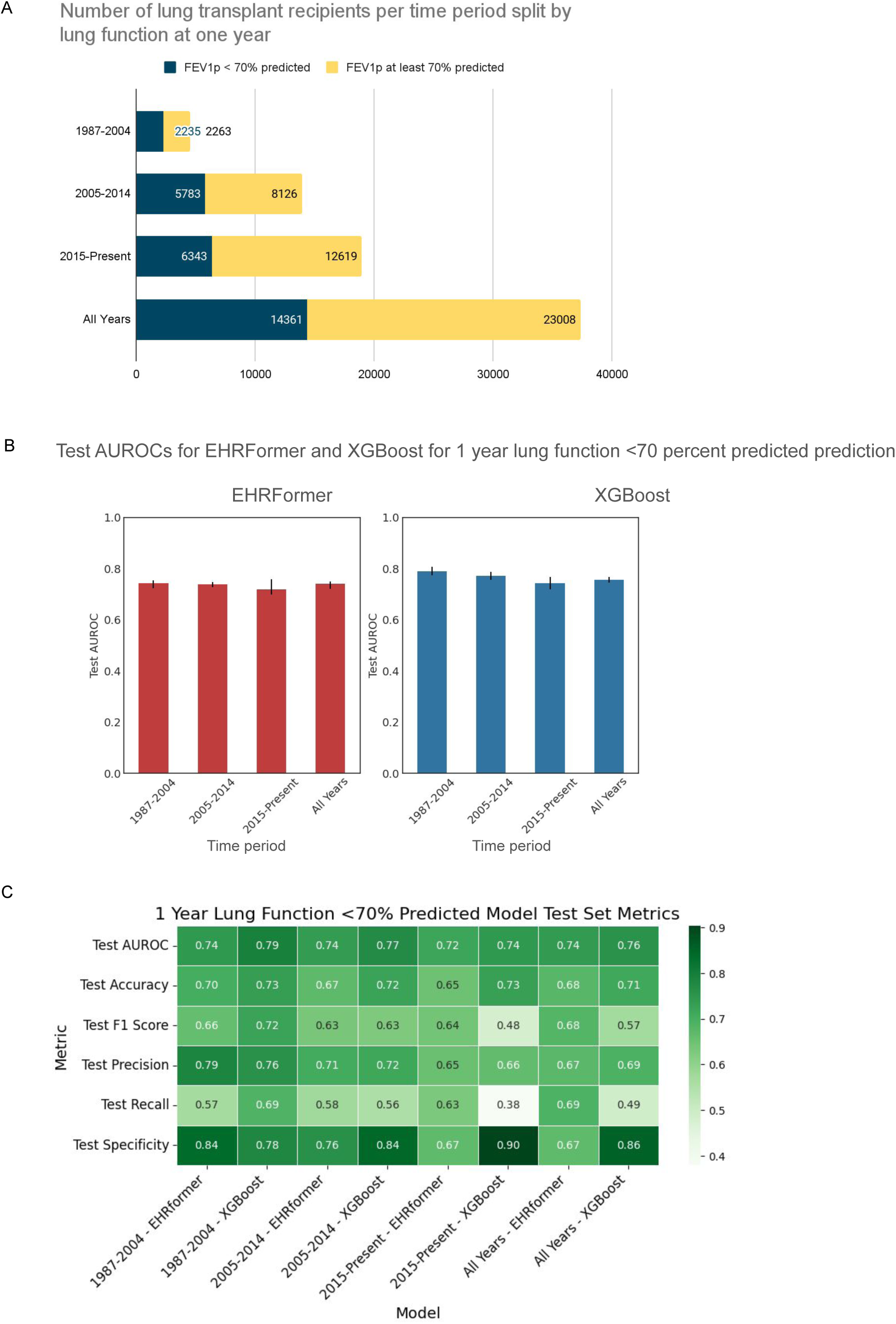
EHRFormer and XGBoost predict one year lung function with strong performance. A. The number of patients in each model split by class outcome of FEV1 <70% predicted at 1 year. B. Test set AUROCs for EHRFormer (red) and XGBoost (blue) prediction of FEV1p <70% predicted across the 4 models. Error bars indicate 95% CIs based on 50 bootstraps of the test set. C. Heatmap of all test set metrics normalized within each metric (row-wise) including AUROC, accuracy, F1 score, precision, recall, and specificity for EHRFormer and XGBoost across the 4 models.

**Table 3:**
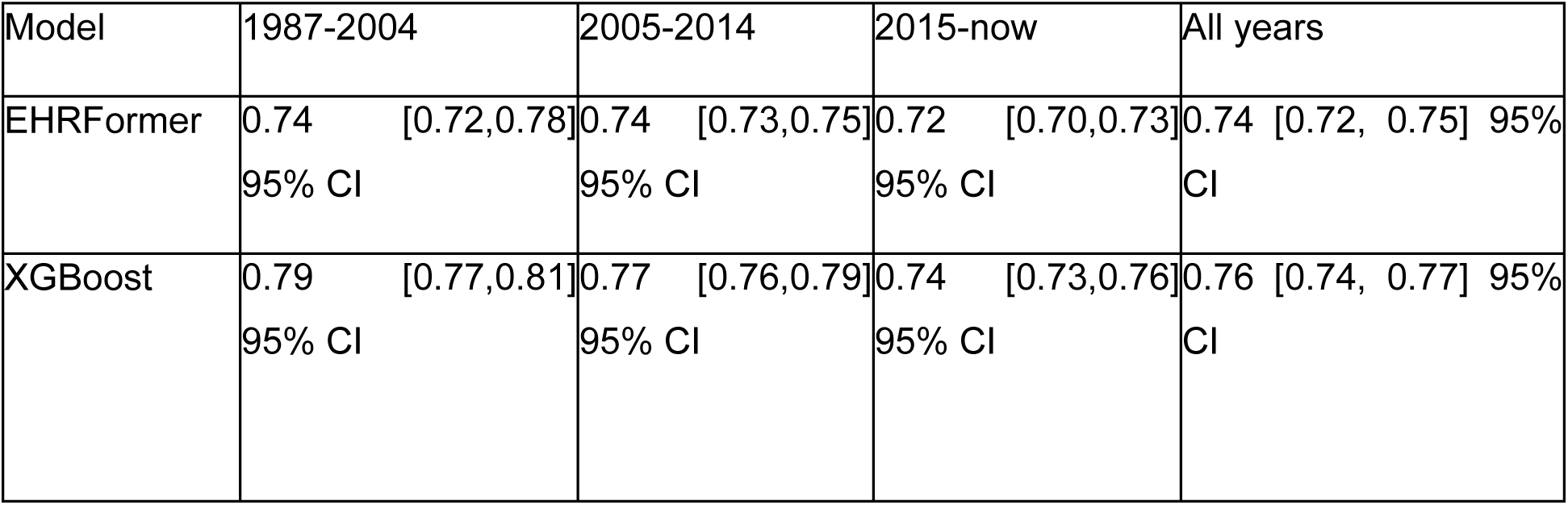
Test AUROCs for EHRFormer and XGBoost at across key time periods in the UNOS dataset.

| Model | 1987-2004 | 2005-2014 | 2015-now | All years |
| --- | --- | --- | --- | --- |
| EHRFormer | 0.74 [0.72,0.78] 95% CI | 0.74 [0.73,0.75] 95% CI | 0.72 [0.70,0.73] 95% CI | 0.74 [0.72, 0.75] 95% CI |
| XGBoost | 0.79 [0.77,0.81] 95% CI | 0.77 [0.76,0.79] 95% CI | 0.74 [0.73,0.76] 95% CI | 0.76 [0.74, 0.77] 95% CI |

### Index stay features have high importance for mortality prediction

To identify features associated with one-year mortality, we developed two XGBoost models. The first used pre-transplant features from all years (Figure 4A, Table S1). The second included both pre-transplant and index hospitalization features (Figure 4B, Table S1). SHapley Additive Probabilities (SHAP) ^22^ values were obtained for both models. Key pre-transplant features for mortality prediction included transplant type, donor ethnicity, PCO2, and FVC at transplant (Figure 4A). For the model including index stay features, important features included length of stay and the occurrence of acute rejection during the index stay (Figure 4B).

**Figure 4:**
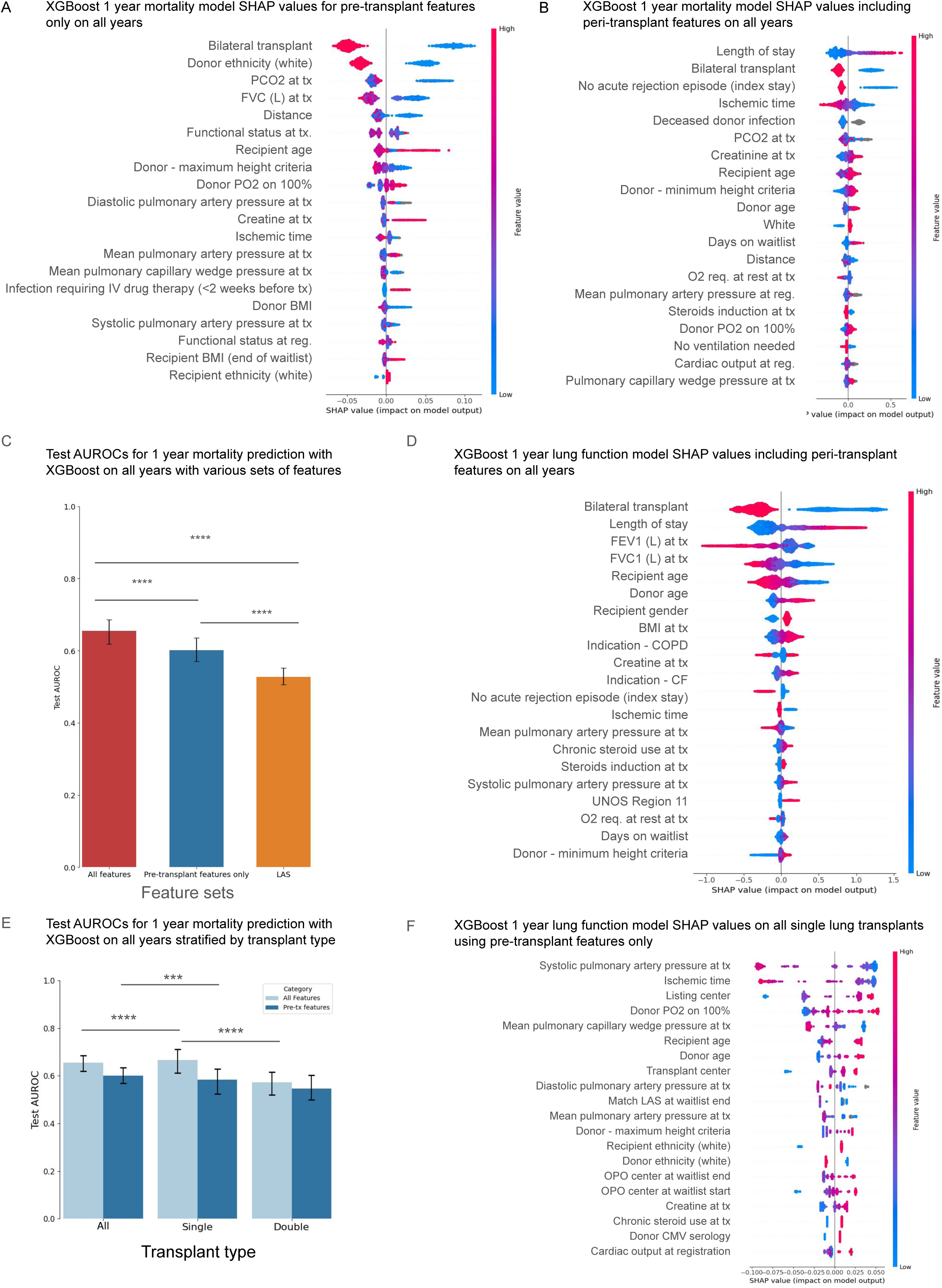
Index stay features strongly influence 1-year outcomes. A. SHAP values from the XGBoost model predicting 1 year mortality using pre-transplant features. B. SHAP values from the XGBoost model predicting 1 year mortality using pre-transplant and peri-transplant features. C. Test AUROCs from XGBoost models predicting 1 year mortality using all features (pre- and peri-transplant), pre-transplant features only, and lung allocation score. Error bars indicate 95% CIs based on 50 bootstraps of the test set. Statistical comparisons were made using the student’s t-test method with correction for FDR <0.05. D. SHAP values from the XGBoost model predicting FEV1p <70% predicted at 1 year using pre- and peri-transplant features. E. Test AUROCs from XGBoost models predicting 1 year mortality by transplant type - all transplants, single, and double lung transplants. Light and dark blue indicate models trained on all features (pre- and peri-transplant features) vs. pre-transplant features only). Highlighted significant statistical comparisons of interest are indicated on the graph. F. SHAP values from the XGBoost model predicting FEV1p <70% predicted at 1 year within single lung transplant recipients using pre-transplant features only.

### Removing index hospitalization features and further subsetting on the Lung Allocation Score (LAS) further reduces model performance

Because index hospitalization features are unavailable when clinicians make decisions to list patients for lung transplantation, we trained another XGBoost model excluding the index stay features specified in Table S1. Model performance for mortality prediction at one year was lower when XGBoost was trained on features only available immediately preceding transplant (Figure 4C and Table 4). Similarly, model performance for mortality prediction at one year was significantly decreased when XGBoost was trained on the initial LAS on the waitlist, the end LAS on the waitlist, the calculated LAS, and the match LAS (Figure 4C and Table 4).

**Table 4:** Test AUROCs for XGBoost for predicting 1 year mortality using all features, pre-transplant features only, and the LAS.

| All features | Pre-transplant features | LAS |
| --- | --- | --- |
| 0.63 [0.60, 0.67] 95% CI | 0.60 [0.57,0.64] 95% CI | 0.53 [0.51,0.55] 95% CI |

### Index stay features are highly influential for prediction of lung function

Important features for predicting lung function included transplant type (single vs. bilateral), ischemic time, PCO2 at registration, creatinine at registration, recipient and donor age, days on the waiting list, and O2 requirement at transplant (Figure 4D and Table 5). . Features unique to lung function prediction were related to pre-transplant lung function (FEV1, FVC), indication type (COPD, cystic fibrosis associated with better function), and recipient BMI (Figure 4D and Table 5).

**Table 5:** Test AUROCs for XGBoost for predicting 1 year mortality using all features or pre-transplant features only stratified by transplant type.

|  | All features | Pre-tx features |
| --- | --- | --- |
| Both | 0.63 [0.60, 0.67] 95% CI | 0.60 [0.57,0.64] 95% CI |
| Single transplants | 0.67 [0.61, 0.71] 95% CI | 0.58 [0.52,0.63] 95% CI |
| Bilateral transplants | 0.57 [0.52, 0.62] 95% CI | 0.55 [0.50,0.60] 95% CI |

### Stratification by transplant type results in a small increase in model performance

After length of stay, transplant type was the most important feature for 1 year mortality. Therefore, we trained separate XGBoost models on all single lung transplant recipients and all double lung transplant recipients. Using all features, including those from the index stay, the test AUROC for single lung transplant recipients was 0.67 [0.61, 0.71], compared to 0.63 [0.60, 0.67] for all patients and 0.57 [0.52, 0.62] for double lung transplant recipients (Figure 4E, Table 5). When modeled on pre-transplant features, subsetting on single lung transplant recipients did not increase model performance compared to modeling on both single and bilateral lung transplants. SHAP analysis within the model trained on single lung transplant recipients using only pre-transplant features revealed the importance of donor and recipient age as well as hemodynamic parameters such as pulmonary arterial pressures. Interestingly, listing center code and center code also emerged as influential features in this model (Figure 4F). Train and test AUROC curves as well as SHAP values for XGBoost models trained on the single and bilateral subsets are shown in Figure S4.

### EHRFormer permits querying the effect of multiple features simultaneously

SHAP values for EHRFormer models are not currently accessible. Instead, we can query feature importance by “perturbing” or changing the value of a specified feature or even a set of multiple features and seeing what effect doing so has on the model’s output. For example, the probability distribution of mortality by one-year does not change after perturbing transplantation region (Figure 5A) but changes substantially after perturbing long index stay (Figure 5B) or dialysis during the index stay (Figure 5C). One can also perform multiple *in silico* perturbations simultaneously. For example, we set positive flags to the highest quartiles for complicated index stay features, including ECMO at 72 hours, inhaled NO at 72 hours, intubation status at 72 hours, and ventilation duration post-transplant which shifted the mortality prediction towards death at one year (Figure 5D).

**Figure 5:**
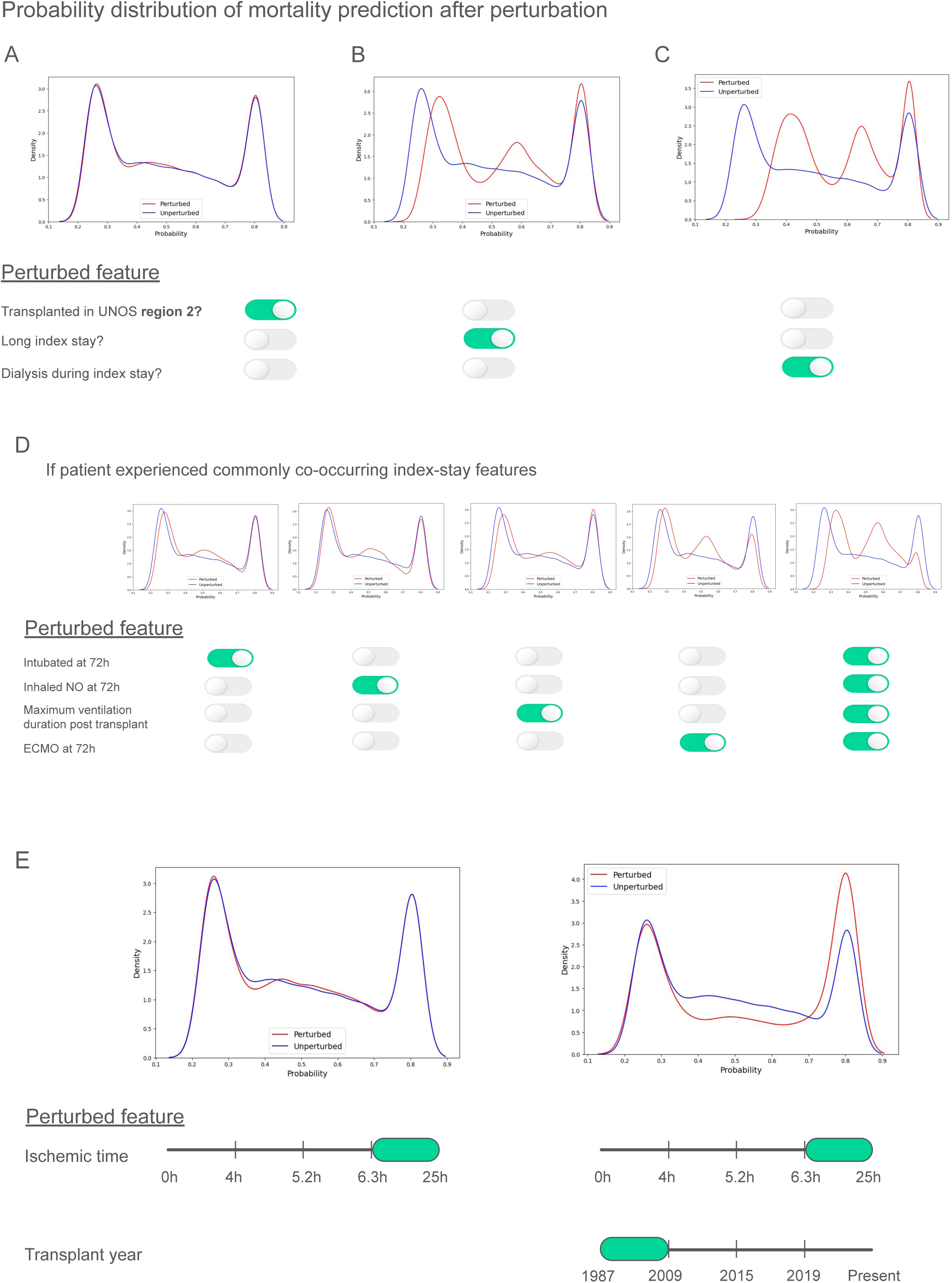
EHRFormer perturbations allow users to query the effect of one or multiple features simultaneously and reveal the paradoxical relationship of ischemic time on mortality. A, B, and C demonstrate probability distribution curves for the model’s prediction of 1 year mortality and how they change when an individual feature is toggled. A. transplantation in UNOS region 2; B. whether patient experienced long index stay; C. whether patient experienced dialysis during index stay. D. Changes in probability distribution curves for 1 year mortality when index stay features reflective of complications are toggled. E. Changes in the probability distribution for 1 year mortality when all observations are set to their maximum quartile for ischemic time are shown on the left. Changes in the probability distribution for 1 year mortality when all observations are set to their maximum quartile for ischemic time and set to their lowest quartile for transplant year are shown on the right.

### Perturbing multiple features simultaneously allows EHRFormer to explain the unexpected influence of long ischemic time on one year mortality in the XGBoost model

Prolonged ischemic time has been historically associated with worse 1-year outcomes ^23^ . Unexpectedly, longer ischemic times were associated with improved outcomes in our XGBoost models (Figure 3A). We hypothesized that the historic association between prolonged ischemic time and poor outcomes were reversed by improvements of organ handling and storage, including the use of *ex-vivo* lung perfusion (EVLP). Accordingly, we leveraged the ability of EHRFormer to query multiple features at once to investigate perturbation of a feature *conditioned on the value of another feature* - in this case what might happen if we prolong ischemic time when transplant year is set to its lowest quartile (earliest) (Figure 5E). When setting ischemic time to its highest value alone, we see a paradoxical shift in mortality prediction that is consistent with the direction of the SHAP values seen in XGBoost (Figure 4A and B). However, when setting ischemic time to its highest value when conditioned on setting transplant year to its lowest quartile, the probability distribution reverses in the opposite direction. Additionally, when we subsetted on those whose lungs underwent EVLP prior to transplant, the proportion of those experiencing poor outcomes such as 1 year mortality, ECMO at 72 hours after transplant, and death during the index stay, were significantly reduced (Figure S6).

### Features associated with frailty predicted death during the index hospitalization

Those who died during the index hospitalization were excluded from our initial mortality prediction models. To investigate features associated with mortality in this subset of patients, we performed hierarchical clustering of all features (Figure 6A). Features associated with mortality during the index stay included transplant indication of idiopathic pulmonary fibrosis or restrictive lung disease, recipients of donors of black or African American ethnicity, and life support features such as ECMO, ventilator, and ICU status. A distinct group of features associated with recipient frailty such as functional status at the time of transplant, infection requiring IV drug therapy prior to transplant, and hospitalization status prior to transplant were associated with higher rates of index hospitalization mortality (Figure 6A). When we investigated features associated with a complicated index hospitalization, those who died during the index stay showed much higher oxygen requirements (FiO2), rates of ECMO, rates of inhaled NO, rates of intubation, and rates of reintubation at 72 hours after transplant (Figure 6B). Similarly, we examined additional frailty features associated with higher rates of index hospitalization mortality. Those who died during the index hospitalization had higher O2 requirements at rest, lower six-minute walk scores, as well as higher rates of chronic steroid use, pan-resistant bacterial infection, infection requiring IV drug therapy, and ventilator status at the time of transplant (Figure 6C).

**Figure 6:**
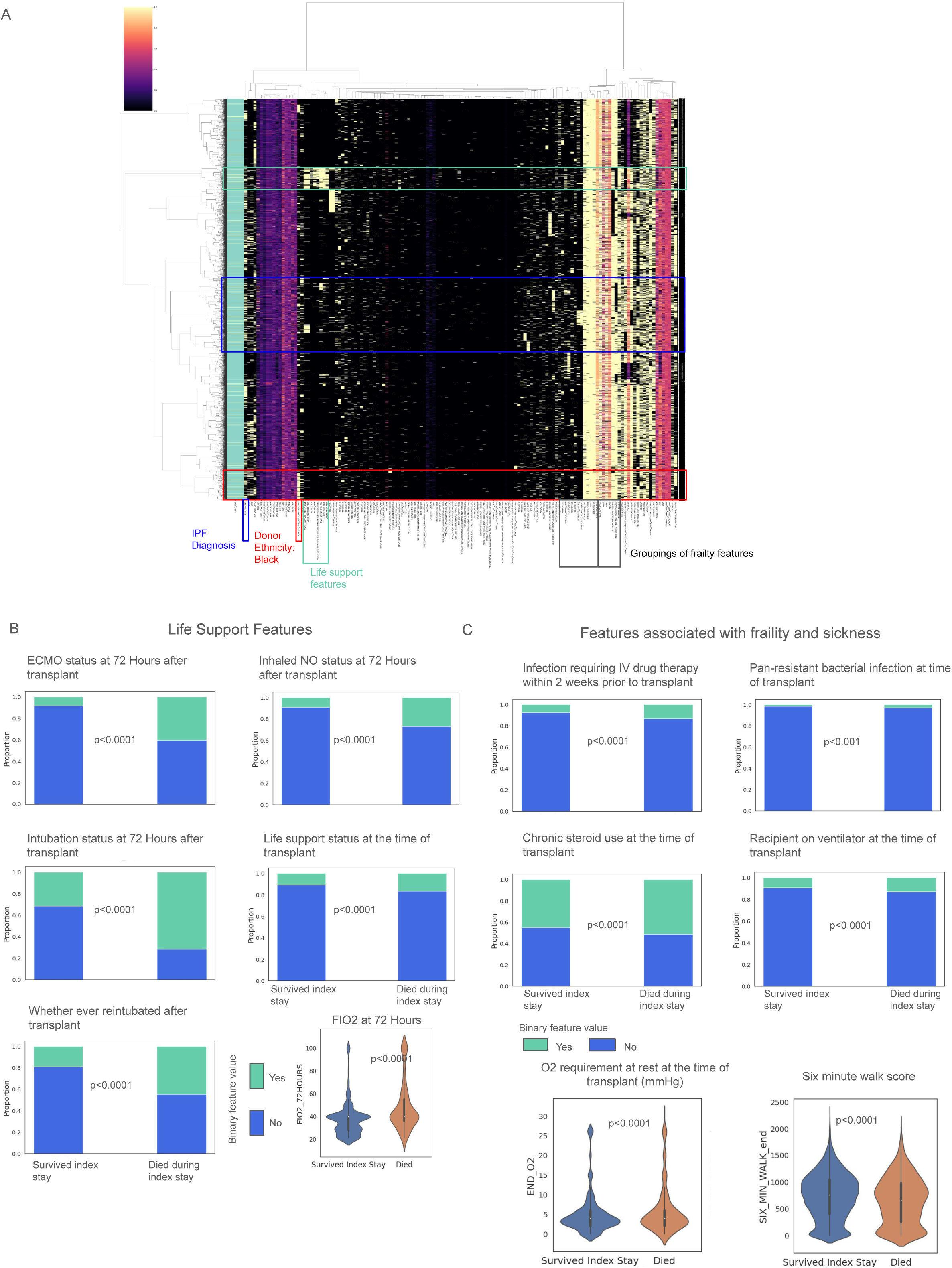
Those who died during their index stay have a higher proportion of flags for life support features, features associated with frailty, and index stay complications. A. heatmap visualization of the hierarchically clustered features used for the prediction tasks. The annotation legend on the left indicates those who died during their index stay (yellow) vs. those who survived the index stay (green). The heatmap also highlights groupings or clusters of features with a higher proportion of those who died during their index stay. Blue box - those whose indication for transplant was idiopathic pulmonary fibrosis; red box - a group of patients highlighted by black donor ethnicity; green box - a set of features associated with life support after transplant; grey boxes - 2 groupings belonging to separate hierarchically clustered “clades” associated with recipient frailty. B. Life support features that were statistically significant in patients who survived the index stay vs. those who died during the index stay, where stacked proportional bar plots were used to represent categorical features (green indicates “yes” and blue indicates “no” for the feature value) and violin plots were used to show continuous features. C. Frailty features that were statistically significant in patients who survived the index stay vs. those who died during the index stay. For graphs in B and C a chi-square (categorical) or Wilcoxon-rank sum (continuous) test was applied with FDR correction <0.05 for multiple comparisons.

## Discussion

Lung transplant is a lifesaving treatment for patients with end stage lung disease. Over the years, lung transplant allocation systems have used prediction models to guide patient eligibility for transplant, organ allocation, and outcome reporting. The role of these models is two-fold: 1) to prioritize organ allocation to patients who have the greatest chance of death due to their underlying lung disease and 2) to direct scarce resources to patients who would achieve maximum survival benefit from lung transplantation. Achieving these goals requires robust prediction models for post-transplant outcomes. Traditionally, these predictive models have been informed by expert-guided supervised selection of features incorporated into traditional statistical methods such as multilinear regression. Modern machine learning approaches such as language models or gradient boosted decision trees can accommodate non-linear relationships between variables and have the potential to account for the multiplicative risks of comorbidities on lung transplant outcomes. We used two robust machine learning models, XGBoost and EHRFormer, which have performed well in other clinical prediction tasks, to predict one-, three- and five-year mortality and lung function after lung transplantation. Our models were trained on data within the UNOS database up until November 2022. Even after optimization, these models performed poorly as predictors of mortality or lung function after transplant, particularly when data from the post-transplant index stay were excluded.

Our study highlights the promise of machine learning approaches in identifying risk factors that drive lung transplant outcomes. We observed a high level of concordance between predictors of poor outcomes identified by our models and those selected by experts for use in supervised models. These included donor and recipient age, length of index stay, and renal dysfunction or failure during the index stay as important predictors of 1 year mortality. Investigation of factors driving our machine learning prediction also revealed biases in the data that would likely be missed in a supervised analysis. For example, the increased risk associated with African American donors likely suggests important risks related to social determinants of health or other factors that would be unlikely to be included in supervised analyses. The failure to include these factors in current models might paradoxically perpetuate these biases in the form of reporting of higher program quality scores for centers in different geographic regions.

XGBoost provides direct measures of the relative weights of features that drive the model. BERT-based models like EHRFormer do not provide explicit information about the factors that drive their predictions. To address this concern, we developed perturbation tools in EHRFormer that allow users to investigate the hypothetical effect of changing variables. We used these to investigate the paradoxical association between prolonged ischemic time and improved mortality after transplantation in the XGBoost models. When we fixed transplant year to the earliest quartile, before procedures such as EVLP were available, increased ischemic time was associated with increased mortality. We show how this tool can be used to perform multiple “perturbations” within EHRFormer simultaneously to query multivariate hypotheses and dependencies. These tools might be helpful to generate hypotheses that can be tested with causal interventions or for individual programs to assess the relative benefits of interventions in the peri-transplant period that can improve outcomes.

Despite their attention to known risk factors associated with transplant outcomes, the performance of our machine learning methods in predicting outcomes was poor. There are several possible reasons that might explain this poor performance. This suggests that data features collected by UNOS are not predictive of transplant outcomes and that collection of additional predictive features such as diffusing capacity of the lungs for carbon monoxide ^24^ is needed. The application of machine learning algorithms to the larger body of EHR data available at individual centers or consortia of centers might identify informative features outside of the UNOS database. Importantly, the models would incorporate molecular features, imaging features, and other modalities outside of EHR data ^25^ . Second, batch effects in the UNOS data related to differences in data curation, collection, reporting between centers, and changes in practice over time might confound the models. Arguing against this, both models identified features previously associated with transplant outcomes as important for their predictions. Applying these models to EHR data that have been validated by clinician review is a strategy to address this concern at the level of individual centers or consortia. Finally, drivers of transplant outcomes might be largely independent of clinical features present before the procedure. The limited performance of these models suggests caution in using these data for predicting lung transplant outcomes.

## Conclusion

In summary, despite their ability to identify and use clinical features known to be associated with lung transplant mortality in their predictions, modern interpretable machine learning approaches applied to the UNOS database performed poorly in predicting one-, three- and five-year survival and lung function after lung transplantation. We developed perturbation tools within EHRFormer to simultaneously explore features in the UNOS database that might reflect changes in lung transplant practice and outcomes over time and unexpected biases in the data. Our data suggest caution when using historic UNOS data to inform clinical practice decisions by multidisciplinary lung transplant teams and outcome reporting. The relative ease with which these models can be applied to more comprehensive clinical and laboratory data, and their demonstrated ability to identify features associated with transplant outcomes, suggest them as powerful approaches to address these limitations.

## Methods

### Data preparation, cleaning, and feature encoding

All observations with a transplant date earlier than November 8, 2022 were included. All waitlist (THORACIC_WL_DATA), peri-transplant (THORACIC_DATA), and follow-up features (THORACIC_FOLLOWUP_DATA) with at least 1 observation were initially considered, yielding 786 features from the STAR File Data Dictionary. All features were reviewed for outliers and unexpected values with consultation from a committee of transplant pulmonologists. Outlier data were treated as missing. Binary features were numerically binarized as 0 or 1. Categorical features were ordinalized if they represented a scale or one-hot encoded. Waitlist and follow-up features were collected at various points in time for each patient. For waitlist data only the first and most recent values were collected. Follow-up observations closest to and within +/-3 months of one-, three-, and five-years post-transplant were used.

### Principal components analysis

The features and resultant dataframe obtained from *Data Preparation and Feature Encoding* were further filtered for those that were missing <10% of data, resulting in 363 features. Numerical features were mean imputed while categorical features were mode imputed. With this dataframe, principal components analysis was applied using *scikit-learn* ^26^ in Python, with the top 50 principal components chosen for initial exploration. *Matplotlib* ^27^ and *seaborn* ^28^ were used for data exploration and visualization.

### Statistical tests

To determine whether continuous variables change with respect to time (transplant date), logistic regression was performed with *statsmodels* ^29^ to determine R-squared coefficients and p-values. To analyze differences in categorical variables between groups, the chi-squared test (*scipy.stats*) ^30^ was used with a Benjamini-Hochberg adjustment for multiple comparisons (FDR < 0.05) *(statsmodels)* ^29^ . To analyze differences in continuous variables between groups, the wilcoxon rank sum test (*scipy.stats*) ^30^ was used with a Benjamini-Hochberg adjustment for multiple comparisons (FDR < 0.05) *(statsmodels*) ^29^ . To draw statistical comparisons between the bootstrapped AUROCs between models (50 bootstraps), a Student’s t-test was performed after assessing normality with the Shapiro-Wilk test (*scipy.stats*) ^30^ . A Benjamini-Hochberg adjustment was made for multiple comparisons (FDR < 0.05) *(statsmodels*) ^29^ .

### Data preparation for modeling

#### Feature selection and preparation for modeling

Subsets of data were chosen for modeling based on observations from the following time frames (inclusive): 1987-2004, 2005-2014, and 2015-Present. These time-related subsets were chosen due to changes in data collection illustrated in S1. All waitlist and peri-transplant data were included for all modeling tasks. Features were filtered for those that were missing <10% of observations within each time period, resulting in 140, 145, 152 and 140 features for data from 1987-2004, 2005-2014, 2015-Present, and all years. Numerical features were mean imputed while categorical features were mode imputed.

#### EHRFormer feature representation and pretraining

From each of the 4 dataframes generated as described in the previous paragraph, 4 different models were pretrained from a tabular BERT initialized with random weights, available via *Huggingface* in the *transformers* package ^31^ . For the base architecture of this model (EHRFormer), we set the standard Transformer Encoder with three transformer layers, with a layer size of 64, an intermediate layer size of 128, and 4 attention heads. For input representation of the tabular data, all numerical values were quantile-binned to represent data from a single patient. Binary features were set to either the lowest or highest quartile bin. Following the standard BERT procedure, we added a 64-dimensional learnable CLS embedding at the beginning of the sequence. An additional bin was included to represent the CLS token for each patient as well as an additional bin for missing data. Each bin and each EHR feature was then assigned with a learnable 64-dimensional vector embedding. We later represented a single EHR entry with a sequence of the length of the number of features, where each element of a sequence was a 64- dimensional vector obtained by summing the feature embedding vector and its assigned bin embedding vector for every feature in the training data. In line with the BERT pretraining procedure, we pre-trained EHRFormer using a masked language model objective. The observations were divided into an 80/20 train/test split for pre-training. During this phase, we randomly masked 15% of the values in the 80% train of each entry by replacing the true bin embedding with a learnable MASK embedding. We later equipped EHRFormer with the task of predicting the actual bin values of the masked entries based on the unmasked EHR feature values. None of the observations in the 20% holdout test set were seen during pretraining.

### Modeling one-, three-, and five-year mortality and lung function outcomes

#### Mortality label retrieval

To obtain the correct labels for one-, three-, and five-year mortality, we used the patient’s date of death (COMPOSITE_DEATH_DATE) to confirm patient death. We then calculated survival in days by subtracting the difference between the patient’s date of death and date of transplant (TX_DATE). Survival time was then used to further identify which patients had died within the one-, three-, and five-year outcomes of interest. To retrieve patients who were alive at one, three, and five years, we used the “patient status”, patient status date, and date of death variables (PX_STAT = “A“ for alive, PX_STAT_DATE, COMPOSITE_DEATH_DATE = NA). PX_STAT_DATE was used to determine total known survival time. We then added this subset of patients to patients who might have died but whose survival exceeded the 1-, 3-, and 5-year outcomes of interest for each task. To further ensure the model was not relying on shortcut features, we removed patients who died during or within 90 days of their index stay. This was determined by filtering out patients whose length of stay (LOS) exceeded survival time by at least 90 days.

#### Lung function <70% of predicted label retrieval

To obtain labels for lung function <70% of predicted, we used the follow-up feature FEV_percent available at 1 year post transplant (+/- 3 months). For patients in which FEV_percent at 1 year post transplant was unavailable, we used the *spiref* ^32^ package with GLI-2012 ^33^ reference values to determine a calculated FEV_percent of predicted based on their absolute FEV1(L) at one year (FEV). Downsampling of the majority class was also applied as described in detail in the subsequent section *EHRFormer fine-tuning and evaluation*.

#### EHRFormer fine-tuning and evaluation

We used fine-tuning of the pretrained EHRFormer for binary classification of the patient’s mortality and lung function at 1 year. Specific outcome retrieval is described above. To perform this task within specific time frames, we used one of the 4 pretrained models that corresponded to these time frames: 1987-2004, 2005-2014, 2015-Present, and all years. With the data and pretrained model from data spanning all years, we also performed identical binary classification of whether the patient was alive or dead at 3 and 5 years. Due to class imbalance for all tasks, the majority class was downsampled at random to result in a 1:1 negative to positive class ratio in both the training and test sets separately.

For each task, following downsampling, hyperparameter tuning was performed using *optuna’s* ^34^ Parzen Tree based estimator (objective = accuracy, direction = maximize, n_trials = 75), on the 80% train split from pretraining. We searched for optimal hyperparameters within the following space: learning_rate = [1e-6, 1e-2], per_device_train_batch_size = [16, 32, 64, 128, 256], and weight_decay = [1e-4, 1e-1]. With the tuned hyperparameters, we performed 5-fold cross-validation on the 80% train split from pretraining. Splits and performance metrics (accuracy, AUROC, F1, precision, recall, and specificity) were determined using *scikit-learn* ^26^ . The tuned models were finally evaluated on their performance using the remaining 20% holdout test set that were not seen during pre-training, hyperparameter tuning, or 5-fold cross-validation. To determine the variability of performance metrics within the test set, 50 random bootstraps were performed on the test set when calculating model metrics.

#### XGBoost evaluation

We similarly used XGBoost for binary classification of the patient’s mortality and lung function at 1 year within time frames corresponding to 1987-2004, 2005-2014, 2015-Present, and all years. We also performed identical binary classification of whether the patient was alive or dead at 3 and 5 years. Additional tasks included assessing the performance of XGBoost on all features, features only available at or before the time of transplant, and on the LAS. For these additional tasks, center-specific features were also included (ie. CTR_CODE, see Table S1). Where center code features were included, the *Catboost Encoder* package was used to perform target encoding of center codes. Finally, XGBoost was also used to determine prediction performance on single and bilateral lung transplant recipients separately. Downsampling of the majority class was done as was done for EHRFormer. Since there is no pre-training process, we divided entire datasets into an 80/20 train/test split at random. 5-fold cross-validation was performed within the training set with *scikit-learn.* Hyperparameter tuning was performed using a Bayesian optimizer with a Gaussian Process based estimator (*scikit-optimize’s BayesSearchCV)* ^35^ . The search space was defined as follows: learning_rate = (0.01, 1.0), max_depth = (2, 12), subsample = (0.1, 1.0), colsample_bytree = (0.1, 1.0), reg_lambda = (1e-9, 100), reg_alpha = (1e-9, 100), min_child_weight = (1, 10), gamma = (0, 5), n_estimators = (50, 1000). To determine the variability of performance metrics within the test set, 50 random bootstraps were performed on the test set when calculating model metrics. For interpretability and insight into XGBoost model decisions, we used SHAP (SHapley Additive exPlanations) ^36^ .

### EHRFormer perturbations

To gain model insights from EHRFormer, we developed a pretrained tabular BERT similar to the pretraining process described in the modeling tasks. We specifically included data from all years and included additional features of interest such as transplant year (TX_YEAR) as well as all the features from the 2015-Present model. The entire dataset was used for pretraining as opposed to setting aside a test cohort. Hyperparameter search was performed within the entire dataset as described previously. We then ran this new model on the fine-tuning binary classification task of 1-year mortality. To understand feature importance, we randomly sampled half of the input observations 10 times. From these sampled observations, we manipulated features of interest by manually changing the bins for one or multiple features in each of the sampled observations. These new “perturbed” inputs were fed to the model during fine-tuning. This process returned new probabilities of a positive class outcome (ie. 1 year mortality) determined by the model when given a perturbed set of features, visualized here as probability distributions before and after the perturbation was applied.

### Hierarchical clustering for length of stay analysis

Hierarchical clustering was performed on all observations using all the features that were used for prediction of 1 year mortality (140 features). Numerical features were mean imputed while categorical features were mode imputed. The ward linkage method and euclidean distance were used. Whether an individual died during their index stay was used as the row annotation feature.

### Propensity score matching for creating matched cohorts for length of stay analysis

To examine associations between those who died during the index stay and those who did not, we performed propensity score matching with psmpy 37 to generate a matched cohort of controls. 1:5 matching was performed based on patients with similar transplant year, indication grouping, gender, age, and ethnicity. Missing data were imputed prior to matching with a simple mean strategy. KNN matching with propensity logits was performed at a 1:5 case:control ratio to mitigate class imbalance.

## Data Availability

All patient-de-identified data was obtained from the United Network for Organ Sharing (UNOS) Standard Transplant Analysis and Research File (SRTR), which is based on the Organ Procurement and Transplantation Network data as of 11/8/2023. Restrictions apply to the availability of these data, which were used under license for the current study, and so are not publicly available. Data are however available from the authors upon reasonable request and with permission from UNOS.

## Disclosure and data availability statement

All patient-de-identified data was obtained from the United Network for Organ Sharing (UNOS) Standard Transplant Analysis and Research File (SRTR), which is based on the Organ Procurement and Transplantation Network data as of 11/8/2023. Restrictions apply to the availability of these data, which were used under license for the current study, and so are not publicly available. Data are however available from the authors upon reasonable request and with permission from UNOS. All research was approved by the Northwestern Institutional Review Board (STU00221316) and all methods were performed in accordance with the relevant guidelines and regulations, including the Declaration of Helsinki. Data were obtained from the UNOS with appropriate permissions. Since UNOS collects data from lung transplant recipients with informed consent, no organs or tissues were procured from prisoners or other vulnerable populations. Data collection for UNOS is conducted under regulations established by the Health Resources and Services Administration (HRSA).

## Code availability

The underlying code for this study is available upon request from the corresponding author at the following repository: https://github.com/NUPulmonary/CLAD_modeling/tree/15f9f50ddc0d8b78ed77e137c5dcc99a12 3b5b12/code/05_UNOS . The training and validation datasets are not publicly available but can be generated from the UNOS database using the code in the previous link.

## Acknowledgements

This research received no funding. This research was supported by the computational resources and staff contributions provided for the Quest high-performance computing facility at Northwestern University, which is jointly supported by the Office of the Provost, the Office for Research, and Northwestern University Information Technology. This research was supported in part through the computational resources and staff contributions provided by the Genomics Compute Cluster which is jointly supported by the Feinberg School of Medicine, the Center for Genetic Medicine, and Feinberg’s Department of Biochemistry and Molecular Genetics, the Office of the Provost, the Office for Research, and Northwestern Information Technology. The Genomics Compute Cluster is part of Quest, Northwestern University’s high-performance computing facility, with the purpose to advance research in genomics. We thank J. Milhans, A. Kinaci, S. Coughlin and all members of the Research Computing and Data Services team at Northwestern for their support.

## Author contributions

LL and MVS conceived of and designed the study. MM, NSM, LL, and AP designed and developed the EHRFormer model. LL, SM, MC, PC, and JL performed data extraction from the UNOS database. LL and MVS conducted data analysis. LL, GRSB, AVM, and MVS wrote the manuscript. LL, MVS, AVM, GRSB, ES, AA, RT, AB, CM, BCB, AB, and AJ provided critical feedback on the manuscript and the analysis described within.

