## Supplementary figures (captions in document) for "*In silico* perturbations provide multivariate interpretability in predicting post-lung transplant outcomes"

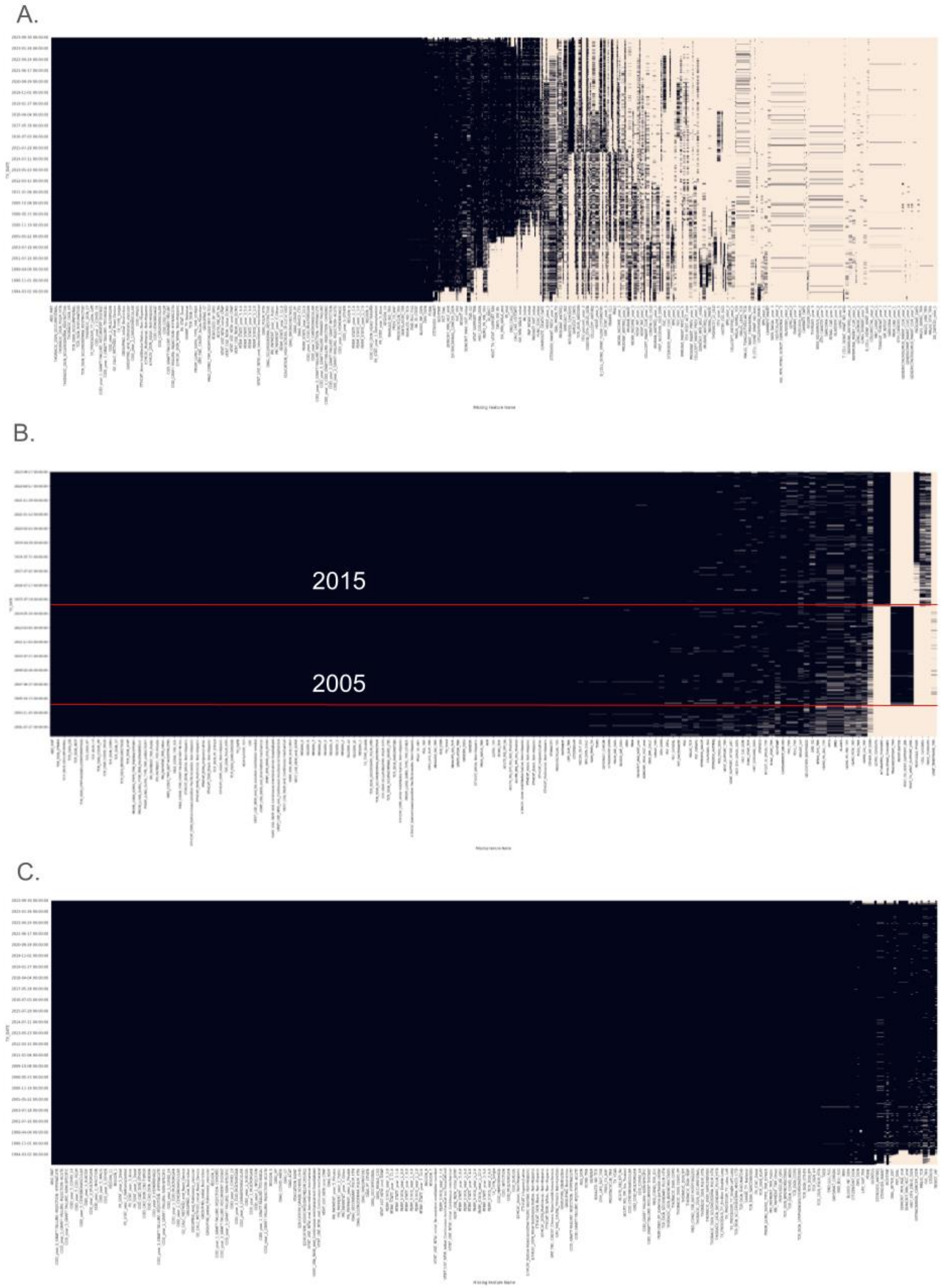

**Figure S1:** Heatmaps showing data missingness of features from Table S1. A. Missingness of all waitlist, follow-up, and peritransplant features sorted by transplant date (780 features). B. Missingness of all waitlist, peritransplant features sorted by transplant date that were at least 90% complete and used for modeling. 2005 and 2015 are indicated on the heatmap to demonstrate features that are missing from the time periods used for modeling. Missingness of all waitlist, follow-up, and peritransplant features sorted by transplant date, used for PCA and at least 90% complete (357 features).

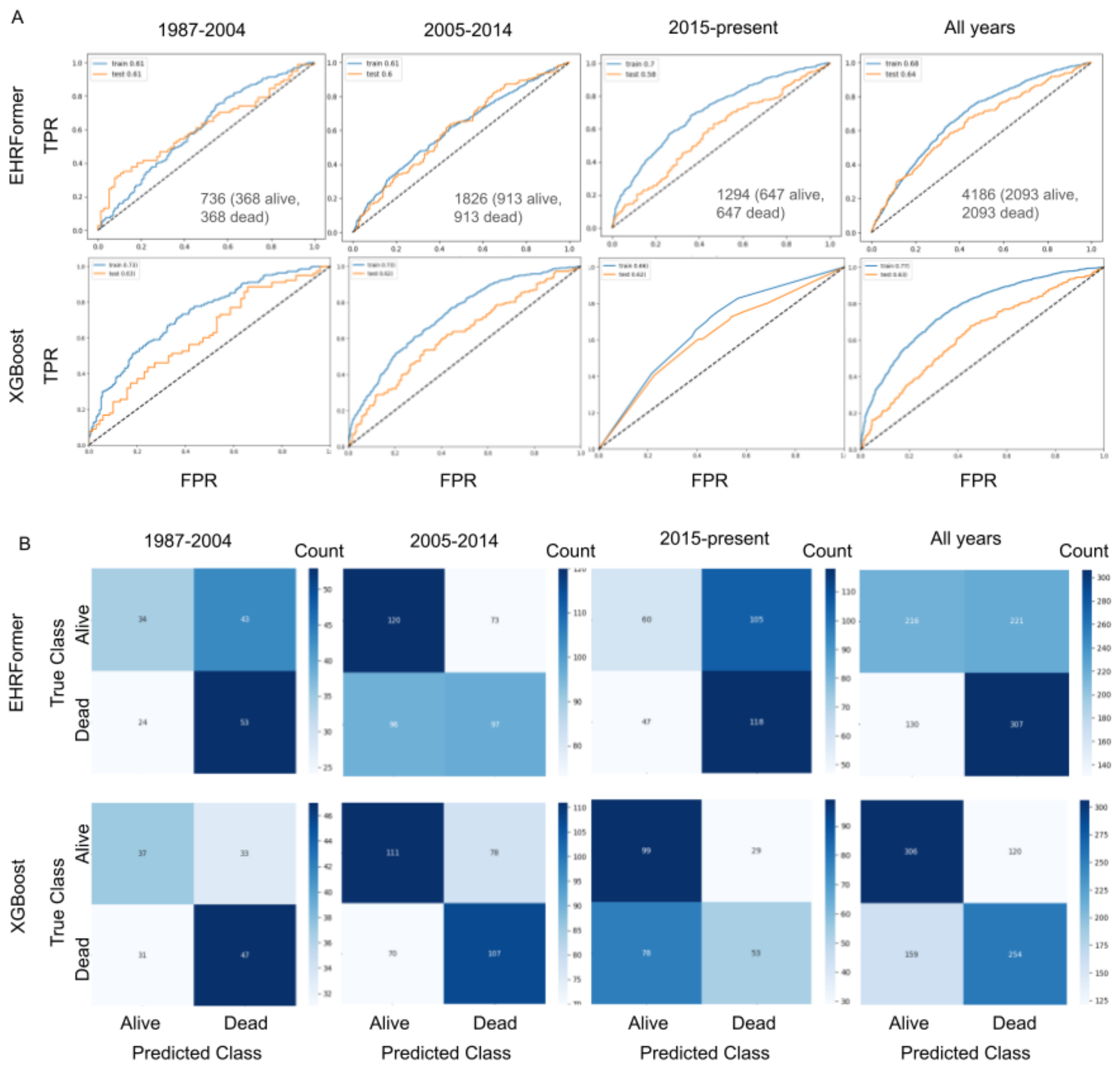


**Figure S2: Area under the receiver operating curves (A) and test set confusion matrices (B) for the prediction of 1 year mortality** using data from 1987-2004, 2005-2014, 2015-present, and from all years. The top row and bottom row in each panel shows the performance in EHRFormer and XGBoost respectively.

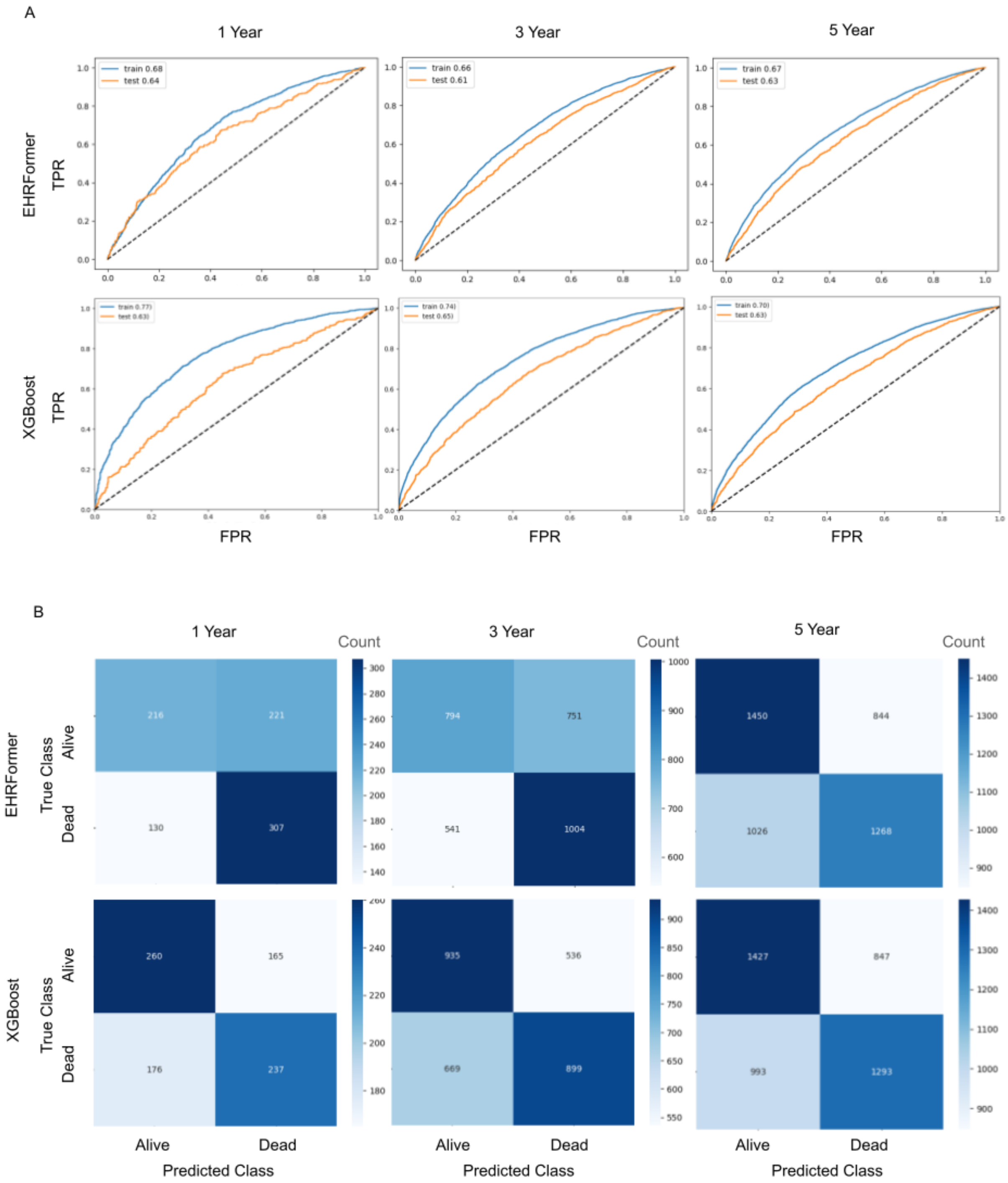


**Figure S3: Area under the receiver operating curves (A) and test set confusion matrices (B) for the prediction of 1, 3, and 5 year mortality** using data from 1987-2004, 2005-2014, 2015-present, and from all years. The top row and bottom row in each panel shows the performance in EHRFormer and XGBoost respectively.

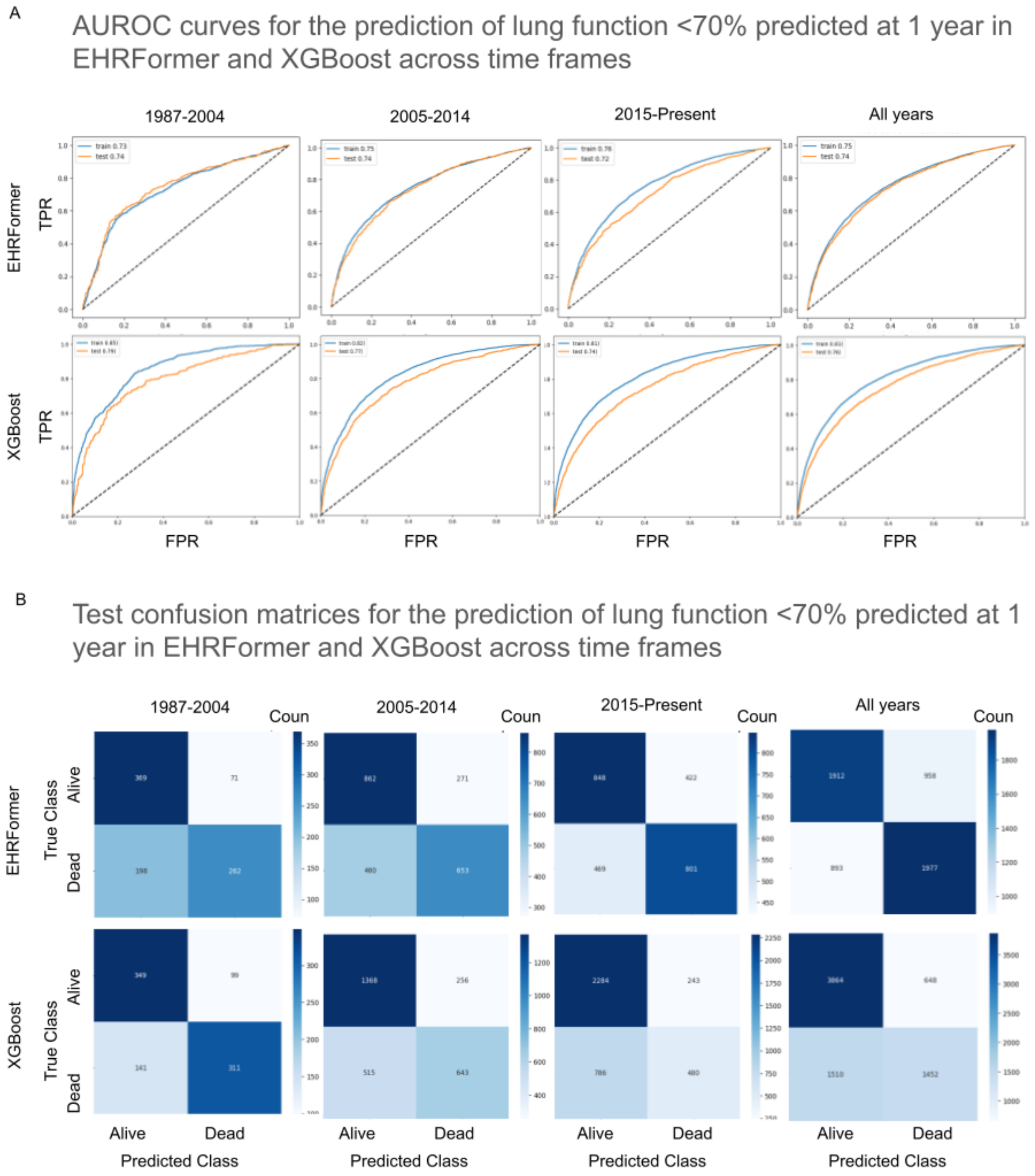


**Figure S4: Area under the receiver operating curves (A) and test set confusion matrices (B) for the prediction of FEV1 <70% at 1 year** using data from 1987-2004, 2005-2014, 2015-present, and from all years. The top row and bottom row in each panel shows the performance in EHRFormer and XGBoost respectively.

A.

AUROC curves for the prediction of 1 year mortality in single vs bilateral lung transplants using XGBoost

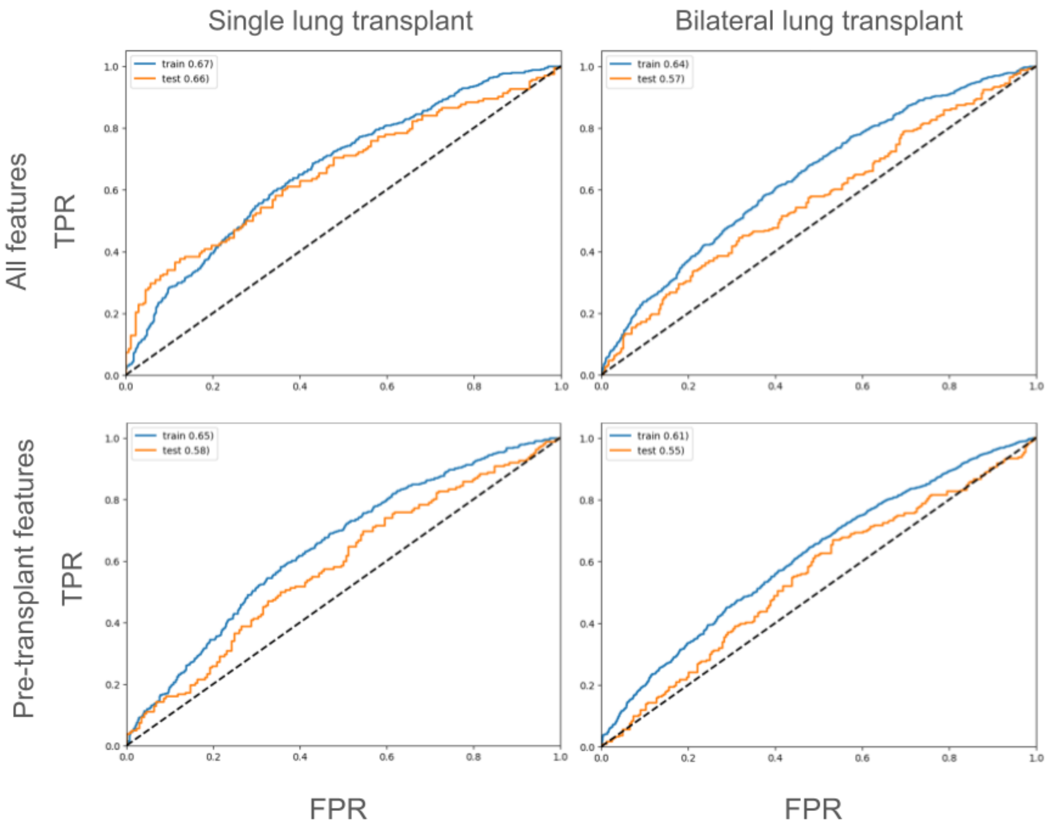


B.

SHAP values for the prediction of 1 year mortality in single vs bilateral lung transplants on **pre-transplant features only** using XGBoost

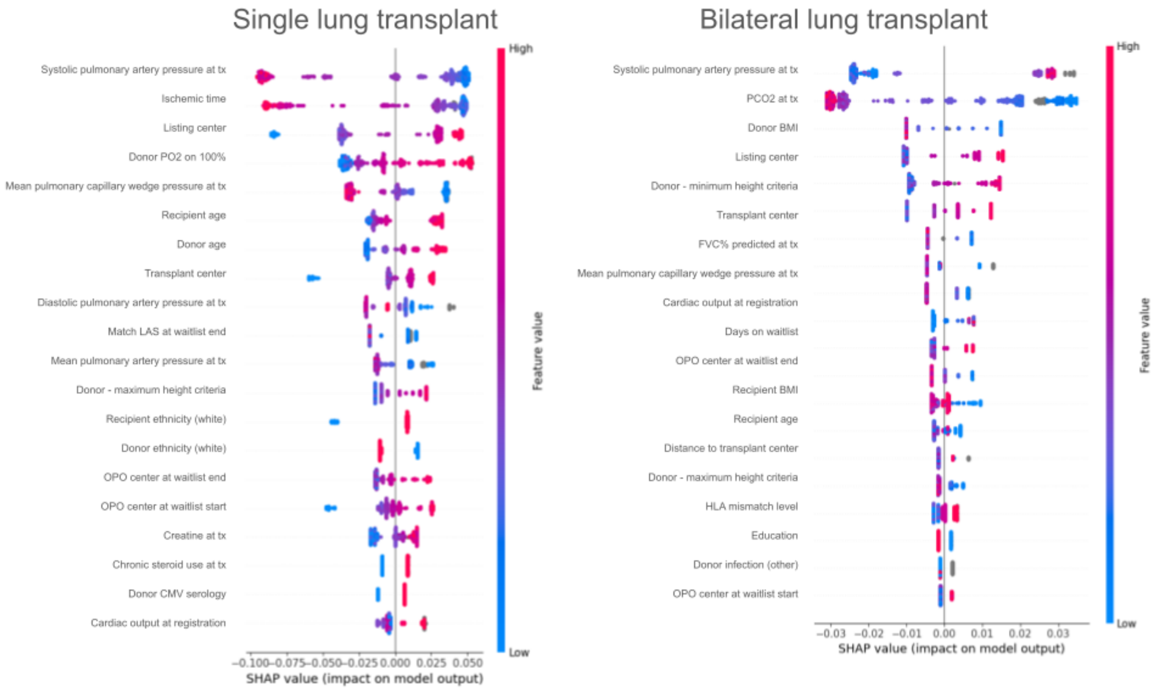

**Figure S5: Area under the receiver operating curves and additional SHAP values for the prediction of 1 year mortality stratified by transplant type using all features vs. pre-transplant features only.** A. Area under the receiver operating curves for XGBoost models predicting 1 year mortality in single vs bilateral lung transplant recipients using either all features or only pre-transplant features as specified in Table S1. B. SHAP values for 1 year mortality prediction single lung transplant (left) vs bilateral lung transplant (right) recipients using only pre-transplant features.

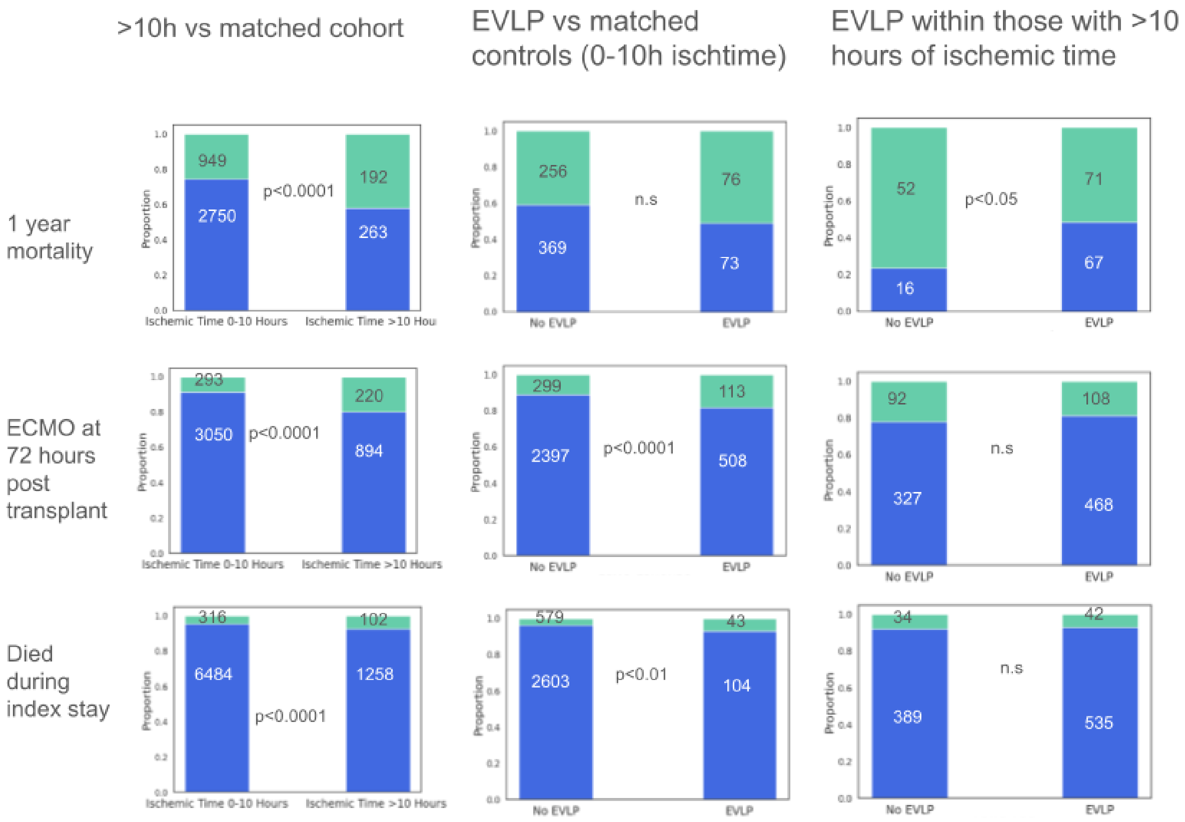


**Figure S6: Stratification of patients by prolonged ischemic time (>10h) reveals a higher proportion of 1 year mortality, ECMO at 72 hours after transplant, and death during the index stay.** Stacked proportional bar plots demonstrating the differences in proportions of 1 year mortality, ECMO at 72 hours after transplant, and death during the index stay (rows) between different stratifications of the patients (columns). Green indicates a positive flag for the binary outcome of interest whereas blue indicates a negative flag. Stratification by EVLP in comparison to matched controls with <10h of lung ischemic time (middle column) revealed some mitigation of poor outcomes. On the other hand, stratification by EVLP in comparison to those with >10h of lung ischemic time (last column) revealed greater mitigation of poor outcomes. Numbers of patients in each grouping are indicated on the graphs.
